# Cardiovascular Outcomes in Low-Risk Non-Diabetic Patients Treated with Semaglutide

**DOI:** 10.1101/2025.05.20.25327768

**Authors:** George Komatsoulis, Alexey Shvets, Zachary Flamholz, Ripple Khera, Samantha Pindak, Nathaniel Tann, Jordan Wolinsky, Anthony Hudzik, Richard Stanford, Manesh Patel, Harry Phillips

## Abstract

Semaglutide use has been shown to reduce cardiovascular events in overweight/obese patients with diabetes and preexisting cardiovascular disease. However, data are limited on the benefit of semaglutide in populations without diabetes and with a low level of preexisting cardiovascular disease. Using claims data from Optum’s de-identified Market Clarity Data, we conducted an observational cohort study to evaluate mortality and cardiovascular outcomes associated with use of semaglutide in overweight/obese patients without diabetes or prior cardiovascular disease (myocardial infarction, stroke, or symptomatic peripheral arterial disease). A total of 38,234 patients who initiated semaglutide between April 2021 and March 2023 were propensity score matched 1:1 to control patients meeting study inclusion and exclusion criteria. Semaglutide patients were treated for a mean (SD) duration of 355 ± 261 days. Semaglutide patients and controls were followed for a mean (SD) duration of 20.2 ± 6.7 and 20.0 ± 6.5 months, respectively. In a time to first event analysis, a composite cardiovascular endpoint (all-cause mortality, nonfatal myocardial infarction or nonfatal ischemic stroke) occurred in 448 (1.2%) semaglutide patients and 645 (1.7%) control patients (HR = 0.73; 95% CI: [0.65, 0.82]; P<0.001). Individual components of the composite endpoint were also significantly lower in the semaglutide group. These findings provide evidence of potential benefit of semaglutide for cardiovascular risk reduction in a population with minimal baseline cardiometabolic complications. Given the population health implications, clinical trials of semaglutide in low risk populations are indicated.

## Background

Overweight and obesity are growing global health challenges with significant economic and health impacts [1,2]. In the United States (US) from 2017-2018, 42.5% of adults were classified as obese (body mass index [BMI] >30) and 31.1% were classified as overweight (BMI 25-30), with obesity rates projected to reach 50% by 2030 [3,4]. In addition to its impact on type 2 diabetes mellitus (T2DM), this obesity epidemic drives significant adverse events and healthcare costs particularly through cardiovascular disease (CVD) which accounts for two-thirds of the 4 million obesity-related yearly deaths and contributes over $250 billion in annual healthcare spending [5,6]. Ultimately, the rising obesity trends in the US and worldwide underscore the growing impact of obesity on the incidence and prevalence of CVD globally [6].

Initially approved to enhance metabolic control in individuals with diabetes, glucagon-like peptide-1 receptor agonists (GLP-1 RAs) have recently demonstrated efficacy for weight loss in adults without diabetes in clinical trials [7–9]. Beyond weight management, GLP-1 RAs have been associated with cardiovascular (CV) benefits, as demonstrated in phase 3 trials (LEADER, SUSTAIN-6, REWIND), reducing the risk of nonfatal myocardial infarction (MI) and stroke in patients with T2DM [10–12]. Unlike these prior studies, SELECT was a multicenter randomized controlled trial that studied the effect of semaglutide on adverse CV outcomes in patients without diabetes who were either overweight or obese and were high-risk for CV events given an inclusion requirement of a history of CVD, including previous MI, previous stroke, symptomatic peripheral arterial disease (PAD), or a combination thereof [13]. In these patients, semaglutide demonstrated a 20% reduction (hazard ratio [HR]: 0.8; 95% confidence interval [CI]: 0.72-0.90; *P*<0.001) in the rate of a composite of death from CV causes, nonfatal MI, or nonfatal stroke vs placebo [13]. While semaglutide has broad approval for weight management and an additional indication for CV risk reduction in established CVD, direct evidence of CV benefit is limited to high-risk populations. The recent CV prevention indication supported by the SELECT trial may improve access for 4.3 million patients with established CVD, but questions remain about benefits in the larger weight management-indicated population without established CVD [14]. The objective of our study was to evaluate CV outcomes in real-world overweight/obese patients without diabetes or prior CVD and inform evidence-based coverage decisions.

## Methods

### Data Source

Optum’s de-identified Market Clarity Data (Optum® Market Clarity) is an integrated, multi-source medical claims, pharmacy claims, and electronic health records data set. Optum® Market Clarity links electronic health record data - including lab results, vital signs and measurements, diagnoses, procedures and information derived from unstructured clinical notes using natural language processing - with historical, linked administrative claim data - including pharmacy claims, physician claims, clinical information facility claims and medications prescribed and administered. Optum® Market Clarity is statistically de-identified under the HIPAA Privacy Rule’s Expert Determination method and managed according to Optum® customer data use agreements [15]. Claims data from April 2020 to March 2024 were used in the analysis.

### Study Design and Study Population

This is an observational longitudinal cohort study that utilized a propensity score matching (PSM) approach to construct comparable cohorts: a semaglutide-treated group and a control group of individuals who did not have a history of ever receiving semaglutide. Patients with a BMI ≥25 determined by International Classification of Disease [ICD]-10-Clinical Modification [CM] codes were split into 2 cohorts: those patients who had ≥1 dispensing of semaglutide between April 2021 and March 2023 and the control group with no exposure to any GLP-1 RAs. Patients in the semaglutide-treated group had a BMI determination within 90 days prior to the initial dispensing date for semaglutide. This dispensing date became the index date for the semaglutide cohort (Supplemental Figure 1). The non-semaglutide-treated control group was assigned random index dates that matched the distribution of the index dates of the semaglutide group, and these dates were within 90 days of a BMI ≥25 ICD-10-CM code. The following exclusions were applied to both treatment and control groups: 1) aged <18 years at index date, 2) ≥1 diagnosis code for diabetes or ≥1 dispensing for a diabetes treatment (except metformin) anytime during the study period (April 2020 to March 2024), 3) <12 months of claims data prior to index date, 4) ≥1 diagnosis code for MI, previous stroke, or symptomatic PAD, 5) ≥1 diagnosis code for any cancer, excluding non-melanoma skin cancer, 6) other weight loss medication use, 7) ≥1 diagnosis code for end stage renal disease, and 8) <12 months of follow-up after index date. The full list of conditions and medications used for the exclusion criteria are reported in Supplemental Table 1. Metformin use was not excluded as the use of metformin without a diabetes diagnosis aligns with the American Diabetes Association’s guidelines for treatment of pre-diabetes [16].

Patients were followed until death, end of claims data, or for a maximum of 3 years whichever came first. Duration of treatment was defined as the time from the initial prescription dispensing to the date of the last prescription dispensing plus 30 days to account for use of the final dispensed product. Because follow-up was terminated at 3 years, any value >1095 days was truncated to 1095. To promote transparency and reproducibility, this study was conducted and reported following the Strengthening the Reporting of Observational Studies in Epidemiology (STROBE) guidelines [17].

Outcomes of interest were defined using ICD-10-CM codes in any position and were captured in the post-index follow-up period. Outcomes included a composite CV endpoint (all-cause mortality or nonfatal MI or nonfatal ischemic stroke [stroke]), and individual outcomes (all-cause mortality, nonfatal MI, and stroke).

### Statistical analysis

PSM was used to achieve balance between the semaglutide and control groups based on measured observable characteristics. Propensity scores were generated on a patient level and were designed to estimate the likelihood of initiating injectable semaglutide. Baseline covariates included in the matching were patient-level demographics (gender, race, age), year of index, comorbid conditions (diagnosis codes and prescriptions) prior to index (heart failure, non-melanoma skin cancer, anticoagulants, antiplatelet drugs, hyperlipidemia, hypertension, mood disorder, albuminuria), mean baseline BMI value, BMI value within 90 days of index (obesity, overweight, and morbid obesity), and smoking history (yes/unknown). Following the determination of propensity scores, a 1:1 nearest-neighbor greedy matching algorithm was utilized to pair each patient on injectable semaglutide with a non-semaglutide-treated control patient who exhibited a similar probability of receiving treatment. The success of this matching process was evaluated through standardized mean differences (SMD), with acceptable thresholds set at <0.1 [18–20].

A Cox proportional hazards model with censoring was used to determine cause-specific HRs and 95% CIs for the first event of the composite CV endpoint which is all-cause mortality, nonfatal MI, and nonfatal ischemic stroke. For the composite CV endpoint, the first event among death, MI, or stroke was included. For the individual endpoints of MI and stroke, an Aalen-Johansen estimator was used where death was considered a competing event to the exposure of interest [21]. The returned cumulative incidence function corresponds to risk of MI and stroke only.

A sensitivity analysis was conducted by excluding patients with high probability of pre-diabetes defined as use of metformin without any diabetes diagnostic codes in the 12 months prior to the index date. A PSM analysis was conducted in this subcohort, wherein semaglutide-treated patients were matched to the non-semaglutide-treated control group as outlined in the primary analysis. All outcomes from the primary analysis were also evaluated in this sensitivity analysis. All analyses were conducted in the Optum Deidentified Data Warehouse environment in python, version 3.9.

## Results

After PSM, the study included a total of 76,468 patients (semaglutide, n=38,234; control, n=38,234) (Supplemental Figure 2); baseline patient characteristics were balanced between the groups, with all SMD for key covariates falling below the predefined threshold of 0.1 (Table 1). Baseline age was comparable between the cohorts, with a mean (SD) age of 46.3 ± 11.5 years for the semaglutide group and a mean (SD) age of 45.9 ± 15.0 years for the control group. Based on the BMI ICD-10-CM code <90 days before index, the semaglutide-treated group and control group were 8.1% and 6.7% overweight, 55.9% and 56.8% obese, and 35.9% and 36.5% morbidly obese, respectively. Healthcare encounter days were similar between study populations in the 12 months prior to index dates, with the semaglutide-treated group having 21.9 ± 21.9 distinct healthcare encounter days compared to 22.5 ± 28.7 distinct healthcare encounter days in the control group. In the follow-up period, the mean (SD) number of filled prescriptions for injectable semaglutide was 8.4 ± 6.6 with a mean (SD) duration of treatment of 355 ± 261 days. The mean (SD) duration of follow-up was 20.2 ± 6.7 months in the semaglutide-treated patients vs 20.0 ± 6.5 months for the control group.

**Table 1.** Baseline Demographics Before and After Matching for Semaglutide and Control Cohorts.

|  | Before propensity-score matching |  |  |  | After propensity-score matching |  |  |  |
| --- | --- | --- | --- | --- | --- | --- | --- | --- |
| Characteristic | Overall<br>(n=543217) | Semaglutide<br>(n=38247) | Control<br>(n=504970) | Std<br>Diff | Overall<br>(n=76468) | Semaglutide<br>(n=38234) | Control<br>(n=38234) | Std<br>Diff |
| Demographic |  |  |  |  |  |  |  |  |
| Age, mean (SD) | 49.7 ± 15.6 | 46.3 ± 11.5 | 49.9 ± 15.8 | 0.265 | 46.1 ± 13.4 | 46.3 ± 11.5 | 45.9 ± 15.0 | 0.025 |
| Male, No. (%) | 202872<br>(37.3) | 7622 (19.9) | 195250<br>(38.7) | 0.421 | 14794<br>(19.3) | 7622 (19.9) | 7172<br>(18.8) | 0.03 |
| Female, No. (%) | 340304<br>(62.6) | 30621 (80.1) | 309683<br>(61.3) | 0.421 | 61668<br>(80.6) | 30608 (80.1) | 31060<br>(81.2) | 0.03 |
| Caucasian, No. (%) | 374939<br>(69.0) | 26021 (68.0) | 348918<br>(69.1) | 0.023 | 52082<br>(68.1) | 26011 (68.0) | 26071<br>(68.2) | 0.003 |
| African American,<br>No. (%) | 69788<br>(12.8) | 3817 (10.0) | 65971<br>(13.1) | 0.097 | 7578 (9.9) | 3817 (10.0) | 3761 (9.8) | 0.005 |
| Asian, No. (%) | 7444 (1.4) | 377 (1.0) | 7067 (1.4) | 0.038 | 754 (1.0) | 377 (1.0) | 377 (1.0) | 0 |
| Other/Unknown,<br>No. (%) | 91046<br>(16.8) | 8032 (21.0) | 83014<br>(16.4) | 0.117 | 16054<br>(21.0) | 8029 (21.0) | 8025<br>(21.0) | 0 |
| Administrative, mean (SD) |  |  |  |  |  |  |  |  |
| No. filled<br>prescriptions for<br>semaglutide | NA | 8.4 ± 6.6 | NA | NA | NA | 8.4 ± 6.6 | NA | NA |
| Days on therapy | NA | NA | NA | NA | NA | 355 ± 261 | NA | NA |
| Follow up medical<br>claims (months) | 20.6 ± 6.4 | 20.2 ± 6.7 | 20.7 ± 6.4 | 0.071 | 20.1 ± 6.6 | 20.2 ± 6.7 | 20.0 ± 6.5 | 0.03 |
| Follow up Rx claims<br>(months) | 20.8 ± 6.9 | 20.2 ± 7.2 | 20.9 ± 6.9 | 0.092 | 20.1 ± 7.0 | 20.2 ± 7.2 | 20.1 ± 6.9 | 0.025 |
| Days with filed<br>medical claim in year<br>prior to index | NA | NA | NA | NA | NA | 21.9 ± 21.9 | 22.5 ± 28.7 | 0.024 |
| Year of Therapy Initiation, No. (%) |  |  |  |  |  |  |  |  |
| 2020 | 12298 (2.3) | 923 (2.4) | 11375 (2.3) | 0.011 | 1697 (2.2) | 921 (2.4) | 776 (2.0) | 0.026 |
| 2021 | 134001<br>(24.7) | 8123 (21.2) | 125878<br>(24.9) | 0.088 | 15877<br>(20.8) | 8122 (21.2) | 7755<br>(20.3) | 0.024 |
| 2022 | 305623<br>(56.3) | 19685 (51.5) | 285938<br>(56.6) | 0.104 | 39419<br>(51.5) | 19683 (51.5) | 19736<br>(51.6) | 0.003 |
| 2023 | 91295<br>(16.8) | 9516 (24.9) | 81779<br>(16.2) | 0.216 | 19475<br>(25.5) | 9508 (24.9) | 9967<br>(26.1) | 0.028 |
| BMI and Overweight/Obesity |  |  |  |  |  |  |  |  |
| BMI<sup>a</sup>, mean (SD) | 34.0 ± 6.0 | 37.6 ± 5.5 | 33.7 ± 6.0 | 0.672 | 37.7 ± 5.8 | 37.6 ± 5.5 | 37.8 ± 6.1 | 0.034 |
| Overweight, No. (%) | 171792<br>(31.6) | 3110 (8.1) | 168682<br>(33.4) | 0.656 | 5689 (7.4) | 3110 (8.1) | 2579 (6.7) | 0.053 |
| Obesity, No. (%) | 266916<br>(49.1) | 21382 (55.9) | 245534<br>(48.6) | 0.146 | 43095<br>(56.4) | 21382 (55.9) | 21713<br>(56.8) | 0.017 |
| Morbid obesity, No. (%) | 104509<br>(19.2) | 13755 (36.0) | 90754<br>(18.0) | 0.414 | 27684<br>(36.2) | 13742 (35.9) | 13942<br>(36.5) | 0.011 |
| Smoking, No. (%) |  |  |  |  |  |  |  |  |
| Current - Yes | 78283<br>(14.4) | 3021 (7.9) | 75262<br>(14.9) | 0.222 | 5898 (7.7) | 3021 (7.9) | 2877 (7.5) | 0.014 |
| Current - Unknown | 464934<br>(85.6) | 35226 (92.1) | 429708<br>(85.1) | 0.222 | 70570<br>(92.3) | 35213 (92.1) | 35357<br>(92.5) | 0.014 |
| Ever - Yes | 103624<br>(19.1) | 4491 (11.7) | 99133<br>(19.6) | 0.218 | 8805<br>(11.5) | 4490 (11.7) | 4315<br>(11.3) | 0.014 |
| Ever - Unknown | 439593<br>(80.9) | 33756 (88.3) | 405837<br>(80.4) | 0.218 | 67663<br>(88.5) | 33744 (88.3) | 33919<br>(88.7) | 0.014 |
| Comorbidities [n (%)] |  |  |  |  |  |  |  |  |
| Heart failure | 15654(2.9) | 568 (1.5) | 15086 (3.0) | 0.102 | 1127 (1.5) | 568 (1.5) | 559 (1.5) | 0.002 |
| Non-melanoma skin cancer | 9031 (1.7) | 431 (1.1) | 8600 (1.7) | 0.049 | 844 (1.1) | 431 (1.1) | 413 (1.1) | 0.005 |
| Mood disorder | 131084<br>(24.1) | 11245 (29.4) | 119839<br>(23.7) | 0.129 | 22422<br>(29.3) | 11238 (29.4) | 11184<br>(29.3) | 0.003 |
| Hyperlipidemia | 234919<br>(43.2) | 15784 (41.3) | 219135<br>(43.4) | 0.043 | 31006<br>(40.5) | 15777 (41.3) | 15229<br>(39.8) | 0.029 |
| Hypertension | 241361<br>(44.4) | 15643 (40.9) | 225718<br>(44.7) | 0.077 | 30826<br>(40.3) | 15637 (40.9) | 15189<br>(39.7) | 0.024 |
| CKD | 25655 (4.7) | 812 (2.1) | 24843 (4.9) | 0.152 | 1572 (2.1) | 812 (2.1) | 760 (2.0) | 0.01 |
| Pancreatitis (acute) | 1789 (0.3) | 50 (0.1) | 1739 (0.3) | 0.044 | 96 (0.1) | 50 (0.1) | 46 (0.1) | 0.003 |
| Pancreatitis (chronic) | 716 (0.1) | 14 (0.0) | 702 (0.1) | 0.035 | 24 (0.0) | 14 (0.0) | 10 (0.0) | 0.006 |
| Albuminuria/<br>MA | 4516 (0.8) | 284 (0.7) | 4232 (0.8) | 0.011 | 588 (0.8) | 283 (0.7) | 305 (0.8) | 0.007 |
| COVID-19 | 88159<br>(16.2) | 7007 (18.3) | 81152<br>(16.1) | 0.06 | 14076<br>(18.4) | 7004 (18.3) | 7072<br>(18.5) | 0.005 |
| Medications, No. (%) |  |  |  |  |  |  |  |  |
| ACEi | 74464<br>(13.7) | 4539 (11.9) | 69925<br>(13.8) | 0.059 | 8965<br>(11.7) | 4538 (11.9) | 4427<br>(11.6) | 0.009 |
| ARB | 69413<br>(12.8) | 5358 (14.0) | 64055<br>(12.7) | 0.039 | 10451<br>(13.7) | 5352 (14.0) | 5099<br>(13.3) | 0.019 |
| Beta blocker | 92072<br>(16.9) | 5831 (15.2) | 86241<br>(17.1) | 0.05 | 11438<br>(15.0) | 5828 (15.2) | 5610<br>(14.7) | 0.016 |
| Thiazides | 76151<br>(14.0) | 6323 (16.5) | 69828<br>(13.8) | 0.075 | 12444<br>(16.3) | 6319 (16.5) | 6125<br>(16.0) | 0.014 |
| Dihydropyridine<br>Ca2+ channel blocker | 67863<br>(12.5) | 3890 (10.2) | 63973<br>(12.7) | 0.079 | 7703<br>(10.1) | 3889 (10.2) | 3814<br>(10.0) | 0.007 |
| SSRI | 123755<br>(22.8) | 11952 (31.2) | 111803<br>(22.1) | 0.207 | 23758<br>(31.1) | 11942 (31.2) | 11816<br>(30.9) | 0.007 |
| SNRI | 38821 (7.1) | 4308 (11.3) | 34513 (6.8) | 0.155 | 8534<br>(11.2) | 4304 (11.3) | 4230<br>(11.1) | 0.006 |
| Benzodiazepine | 68489<br>(12.6) | 6753 (17.7) | 61736<br>(12.2) | 0.153 | 13399<br>(17.5) | 6744 (17.6) | 6655<br>(17.4) | 0.006 |
| Atypical<br>antipsychotic | 25745 (4.7) | 1955 (5.1) | 23790 (4.7) | 0.019 | 3916 (5.1) | 1955 (5.1) | 1961 (5.1) | 0.001 |
| Statin | 128304<br>(23.6) | 7157 (18.7) | 121147<br>(24.0) | 0.129 | 13988<br>(18.3) | 7154 (18.7) | 6834<br>(17.9) | 0.022 |
| Aspirin | 13948 (2.6) | 608 (1.6) | 13340 (2.6) | 0.073 | 1180 (1.5) | 608 (1.6) | 572 (1.5) | 0.008 |
| P2Y12i | 6878 (1.3) | 232 (0.6) | 6646 (1.3) | 0.073 | 419 (0.5) | 231 (0.6) | 188 (0.5) | 0.015 |
| Vitamin K<br>antagonist | 4349 (0.8) | 164 (0.4) | 4185 (0.8) | 0.051 | 321 (0.4) | 164 (0.4) | 157 (0.4) | 0.003 |
| Oral anticoagulant | 19642 (3.6) | 939 (2.5) | 18703 (3.7) | 0.072 | 1844 (2.4) | 939 (2.5) | 905 (2.4) | 0.006 |
| Ezetimibe | 7165 (1.3) | 442 (1.2) | 6723 (1.3) | 0.016 | 864 (1.1) | 441 (1.2) | 423 (1.1) | 0.004 |
| Fibrate | 7239 (1.3) | 433 (1.1) | 6806 (1.3) | 0.019 | 866 (1.1) | 433 (1.1) | 433 (1.1) | 0 |
| PCSK9i | 1113 (0.2) | 100 (0.3) | 1013 (0.2) | 0.013 | 193 (0.3) | 99 (0.3) | 94 (0.2) | 0.003 |
| Metformin | 16599 (3.1) | 7004 (18.3) | 9595 (1.9) | 0.566 | 13177<br>(17.2) | 6991 (18.3) | 6186<br>(16.2) | 0.056 |

Abbreviations: ACEi, angiotensin-converting enzyme inhibitor; ARB, angiotensin receptor blocker; BMI, body mass index; CKD, chronic kidney disease; COVID-19, coronavirus disease 2019; MA, microalbuminuria; NA, not applicable; PCSK9i, proprotein convertase subtilisin/kexin type 9 inhibitor; P2Y12i, purinergic receptor type Y subtype 12 inhibitor; SD, standard deviation; Std Diff, standardized mean difference; SNRI, serotonin-norepinephrine reuptake inhibitor; SSRI, selective serotonin reuptake inhibitor.

In a time to first event analysis over 3 years, the composite CV endpoint (all-cause mortality, MI, or stroke) occurred in 448 (1.2%) of 38,234 patients in the semaglutide-treated patients vs 645 (1.7%) of 38,234 in the control group (HR 0.73; 95% CI:0.65-0.82; P<0.001; Table 2).

**Table 2.**
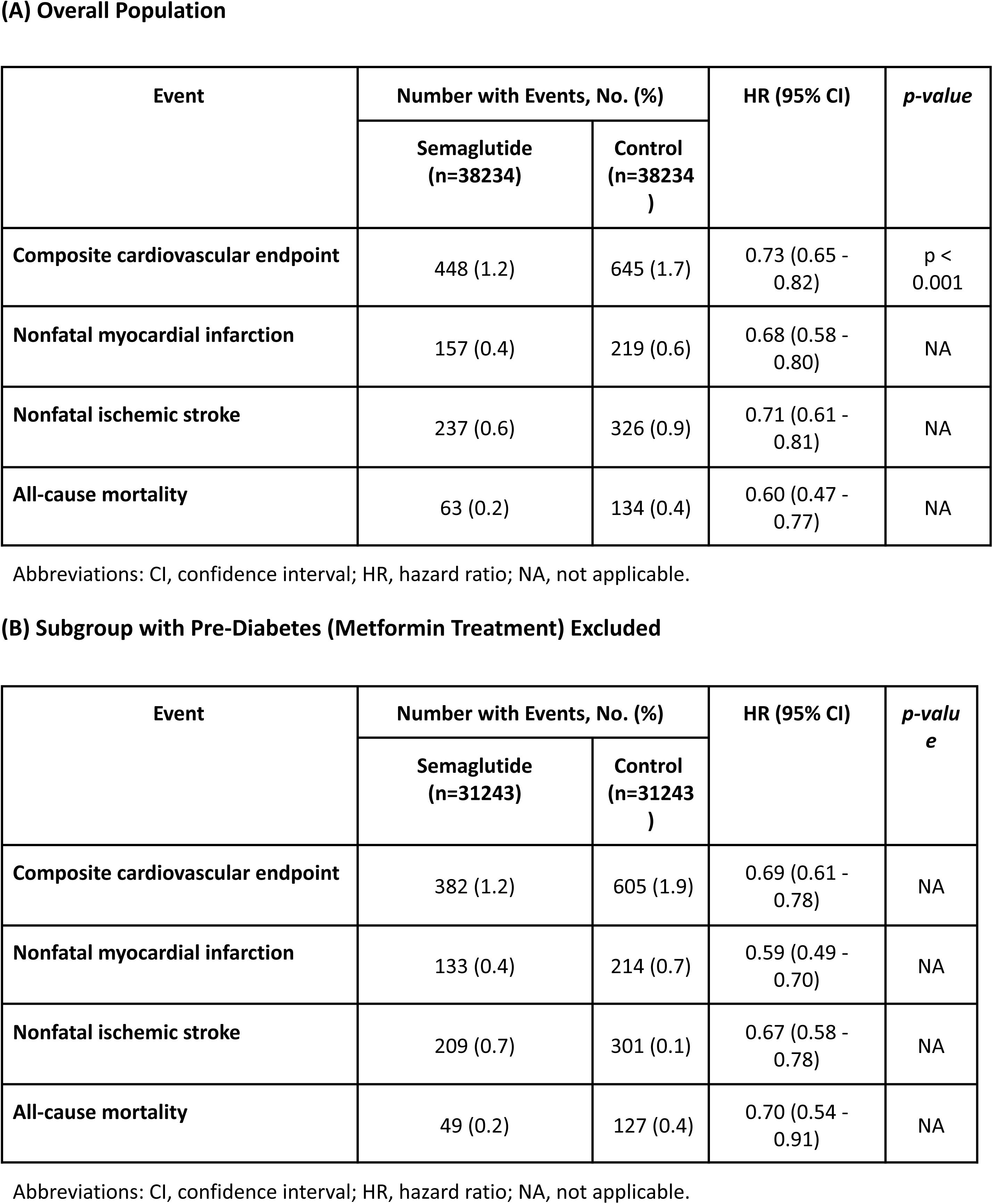
Time-to-First Event Endpoints.

**(A) Overall Population**
| Event | Number with Events, No. (%) |  | HR (95% CI) | *p-value* |
| --- | --- | --- | --- | --- |
|  | Semaglutide (n=38234) | Control (n=38234) |  |  |
| Composite cardiovascular endpoint | 448 (1.2) | 645 (1.7) | 0.73 (0.65 - 0.82) | p < 0.001 |
| Nonfatal myocardial infarction | 157 (0.4) | 219 (0.6) | 0.68 (0.58 - 0.80) | NA |
| Nonfatal ischemic stroke | 237 (0.6) | 326 (0.9) | 0.71 (0.61 - 0.81) | NA |
| All-cause mortality | 63 (0.2) | 134 (0.4) | 0.60 (0.47 - 0.77) | NA |

Abbreviations: CI, confidence interval; HR, hazard ratio; NA, not applicable.

**(B) Subgroup with Pre-Diabetes (Metformin Treatment) Excluded**
| Event | Number with Events, No. (%) |  | HR (95% CI) | *p-value* |
| --- | --- | --- | --- | --- |
|  | Semaglutide (n=31243) | Control (n=31243) |  |  |
| Composite cardiovascular endpoint | 382 (1.2) | 605 (1.9) | 0.69 (0.61 - 0.78) | NA |
| Nonfatal myocardial infarction | 133 (0.4) | 214 (0.7) | 0.59 (0.49 - 0.70) | NA |
| Nonfatal ischemic stroke | 209 (0.7) | 301 (0.1) | 0.67 (0.58 - 0.78) | NA |
| All-cause mortality | 49 (0.2) | 127 (0.4) | 0.70 (0.54 - 0.91) | NA |

Abbreviations: CI, confidence interval; HR, hazard ratio; NA, not applicable.

All-cause mortality occurred in 63 (0.2%) of 38,234 patients in the semaglutide-treated group vs 134 (0.4%) of 38,234 patients in the control group (HR 0.60; 95% CI: 0.47-0.77). MI occurred in 157 (0.4%) of the semaglutide-treated group and 219 (0.6%) in the control group (HR 0.68; 95% CI: 0.58-0.80). Lastly, stroke occurred in 237 (0.6%) of the semaglutide-treated patients and 326 (0.9%) of the control patients (HR 0.71; 95% CI: 0.61-0.81) as shown in Table 2a. The cumulative incidence curves for all endpoints can be seen in Figure 1. In addition, the absolute risk reduction (ARR) for the composite CV outcome was 0.5% (1.2% vs 1.7% in the treatment and control groups, respectively), which corresponds to an estimated number needed to treat (NNT) of 200. Finally, in the sensitivity analysis excluding patients with metformin use (pre-diabetic patients), results remained consistent with the primary analysis (Figure 2 and Table 2b)

**Figure 1.**
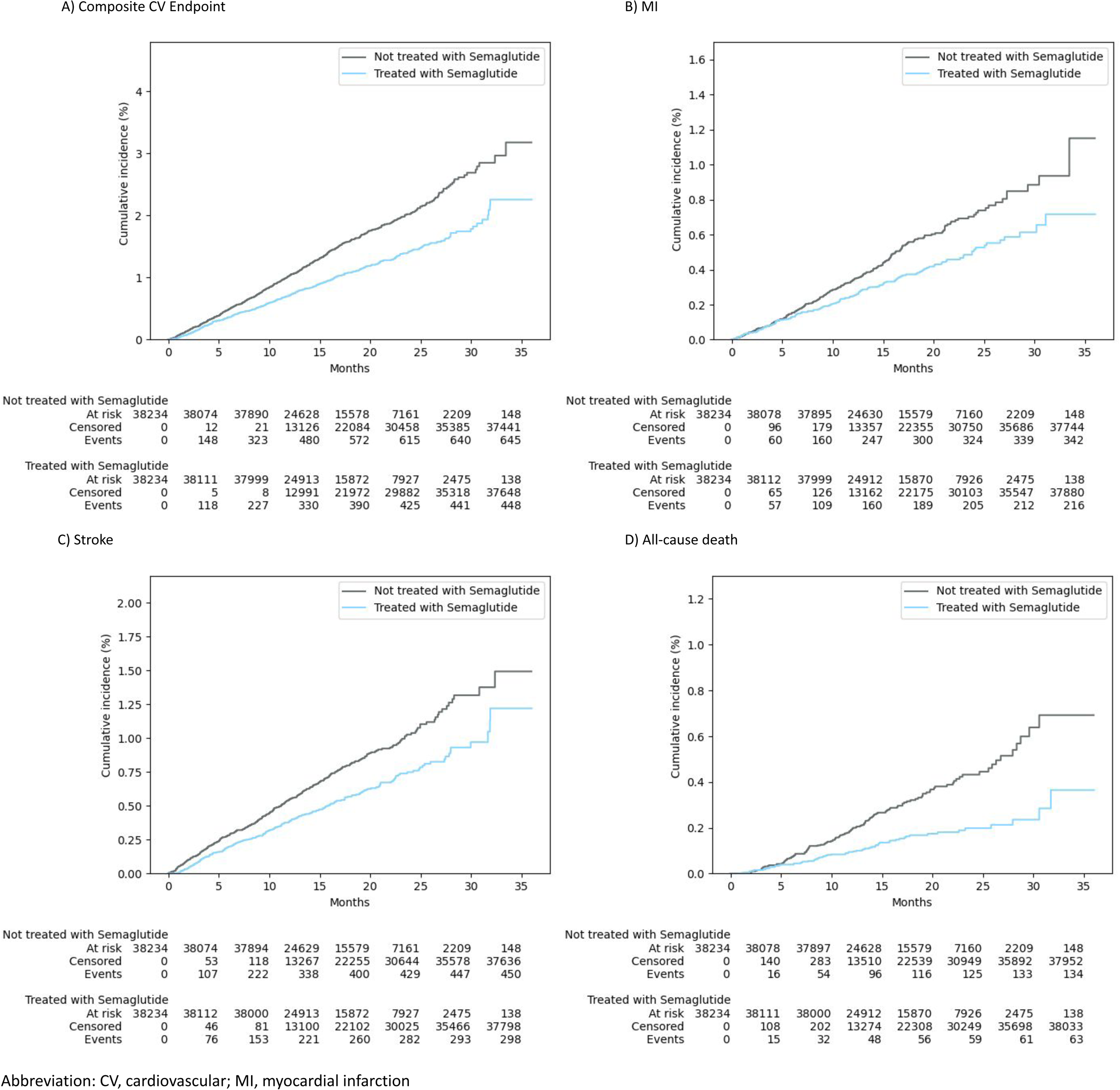
Cumulative Incidence for the Overall Population for All Outcomes.

**Figure 2.**
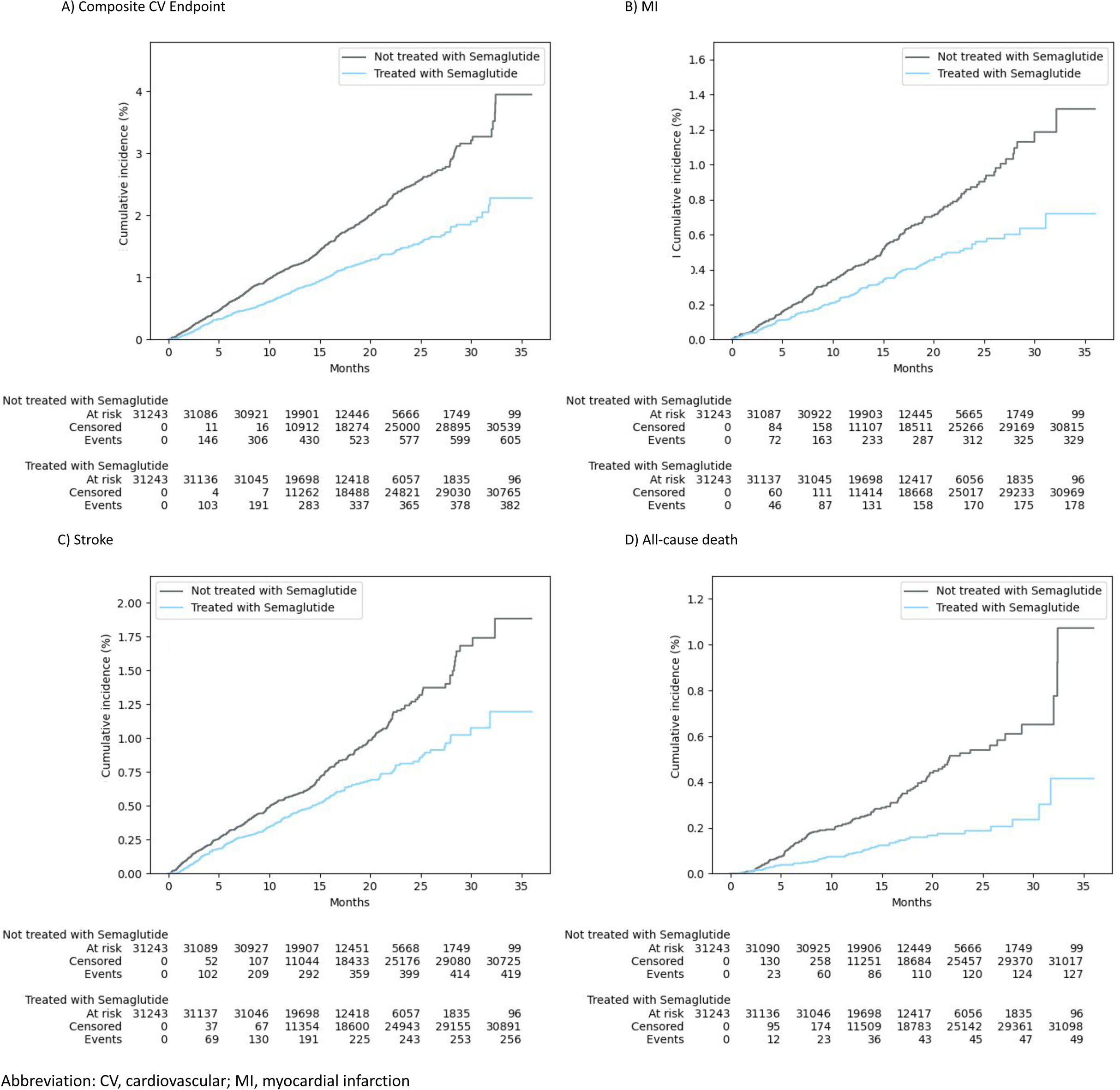
Cumulative Incidence for Subgroup with Pre-diabetes (Metformin Treatment) Excluded.

## Discussion

In this analysis from a large claims database involving adults with overweight or obesity but without diabetes or CVD (MI, stoke, or PAD), we found that patients treated with semaglutide had reduced CV events or mortality compared with matched controls over a mean duration of follow-up of 20.1 months. This association persisted even when patients with pre-diabetes were excluded. To our knowledge, this study is the first to compare the effectiveness of semaglutide with controls in patients eligible for weight management but with minimal cardiometabolic comorbidities. These findings suggest that semaglutide may offer CV risk reduction benefits beyond the approved indications. Expanding the eligible population for semaglutide treatment could have important public health implications by influencing both clinical practice and coverage decisions for CV prevention therapies.

These findings extend the analysis of the SELECT trial to a lower-risk population without preexisting CVD. In SELECT, the CV composite endpoint occurred in 6.5% of semaglutide-treated patients vs 8.0% of placebo-treated patients (HR 0.80; 95% CI: 0.72-0.90) with a 1.5% ARR. In our study, the composite endpoint occurred in 1.2% of semaglutide-treated patients vs 1.7% of the control group (HR 0.73; 95% CI: 0.65-0.82) with a 0.5% ARR. The smaller absolute difference and higher NNT (200 vs 67 in SELECT) is likely a reflection of the lower-risk population without preexisting CVD and shorter follow-up in our analysis.

The mechanism responsible for the significant reduction in CV risk with semaglutide that we observed over a relatively short period of time is unknown, but is likely multifactorial. We observed a divergence in the cumulative incidence curves starting from the initiation of therapy, suggesting that the association cannot be solely attributed to a reduction in body mass. Evidence suggests that GLP-1 RAs may enhance myocardial glucose uptake, thereby contributing to cardioprotective effects and the prevention of adverse cardiac remodeling [22,23]. Additionally, GLP-1 RAs have vasodilatory properties that help lower blood pressure, and they exhibit anti-inflammatory effects, which play a role in reducing the formation and progression of atherosclerotic lesions [24–26]. The anti-inflammatory effect is hypothesized to be the result of GLP-1s multifactorial impact on regulating immunological responses, initiating anti-inflammatory pathways, and decreasing production of pro-inflammatory cytokines [27]. Lastly, GLP-1 RAs impact glycemic control and weight loss which have been associated with CV benefits [13, 28, 29].

Our findings underscore the potential of semaglutide in enhancing CV health within a broader patient demographic. While the higher NNT found in our study compared to SELECT reflects lower baseline risk, the population impact could be substantial given that there is a large lower-risk population potentially eligible for CV risk reduction [14]. There are an estimated 4.3 million US non-diabetic patients who are overweight or obese with CVD who may be eligible for treatment based on SELECT, but our findings suggest potential benefit in a subset of the 97.5 million patients eligible for weight management without diabetes or established CVD [14]. This creates a complex challenge for healthcare systems: balancing individual benefit, population health impact, and resource constraints. Coverage decisions will need to consider both the modest ARR in lower-risk individuals and the potentially large population-level impact. Cost-effectiveness analyses incorporating both individual and population-level benefits will be essential to inform evidence-based coverage decisions.

Cost, inconsistent payer coverage, drug shortages, and side effects have led to short duration of treatment with GLP-1 RAs; patients treated with GLP-1 RAs have lower than 50% persistence rates at 1 year [30]. Despite these real-world adherence challenges, CV benefit was demonstrated in our study including reduced all-cause death even with a modest mean (SD) treatment duration of 355 ± 261 days over a mean duration of follow-up of 20 months. Improving access and support of patients during treatment are important goals in GLP-1 RA management, but the potential benefits of even short durations of treatment should not be discounted, pending further research.

A key strength of our study is the large sample size, which enhances the reliability and generalizability of our findings. We analyzed nearly 28 million patients with BMI ≥25, and derived treatment and control cohorts were more than 4-times larger than those in the SELECT trial [13]. Furthermore, both groups were well-matched following the PSM, which helped minimize bias and enhance the robustness and reliability of the results.

Our study has several limitations. As this is an observational study, we are unable to conclusively establish a causal relationship between the use of semaglutide and the CV results observed. While the PSM effectively aligned both groups, there is still the possibility of unobserved clinical variables which may impact the results. Another limitation is the potential for selection bias, as individuals prescribed semaglutide may differ in characteristics and healthcare seeking behavior compared to those who do not receive this treatment. However, baseline healthcare encounter days (events), using medical claims in the prior 12 months, were balanced between the groups indicating similar healthcare utilization patterns. Furthermore, overweight/obesity were determined by ICD-10-CM codes and not actual BMI values which could lead to misclassification. Even so, the groups were aligned on the broader categories of overweight, obese, and morbidly obese which makes misclassification less likely. In addition, the presence of metformin use without a diabetes diagnosis, which was used as a surrogate for pre-diabetes, may have inadvertently included patients with diabetes in the semaglutide cohort; however, sensitivity analysis excluding this group showed similar results.

In summary, we observed that overweight and obese individuals without diabetes and with low CV risk who were treated with semaglutide had a reduced likelihood of experiencing MI, stroke, or death compared with patients not receiving semaglutide over a 3-year time-period. Although the ARR was smaller than that observed in high-risk populations, the substantial size of the potentially eligible treatment population suggests important public health implications and underscores the need for future clinical studies. Considering that not all patients benefited from treatment and that the event incidences were low, another important next step would be to identify the sub-population or patient profiles most likely to benefit. Integrating additional data modalities and employing cutting-edge technologies that maximize outcome benefits like machine learning, predictive modelling, and artificial intelligence could help to stratify patient benefit from GLP1-RA intervention. These approaches could be utilized at the payer or prescriber level to drive cost-effective treatment decisions.

## Data Availability

All data produced in the present study are available upon reasonable request to the authors.

## Author Contributions

Dr. Komatsoulis had full access to all of the data in the study and takes responsibility for the integrity of the data and the accuracy of the data analysis.

Concept and design: Komatsoulis, Shvets, Flamholz, Hudzik, Stanford, Phillips

Acquisition, analysis or interpretation of data: Komatsoulis, Shvets, Flamholz, Khera, Pindak, Tann, Wolinsky, Hudzik, Stanford, Phillips

Drafting of the manuscript: Komatsoulis, Shvets, Flamholz, Hudzik, Stanford, Phillips

Critical revision of the manuscript for important intellectual content: All authors

Statistical analysis: Komatsoulis, Shvets

Obtained funding: Phillips

Administrative, technical, or material support: All authors

Supervision: Stanford, Phillips

## Data Sharing Statement

The datasets generated during and/or analyzed during the current study are not publicly available

## Additional Contributions

Mayaan Baron provided early statistical support. Candy Zhu provided administrative assistance throughout the execution and publication of this study.

