## Supplemental Material for "Cardiovascular Outcomes in Low-Risk Non-Diabetic Patients Treated with Semaglutide"

**Supplement**

**Content**

Supplemental Table 1. Conditions and Medications Used in the Exclusion Criteria

Supplemental Table 2. BMI Assignment

Supplemental Figure 1. Study Diagram for Semaglutide and Control Cohorts

Supplemental Figure 2. Study Flow Diagram

**Supplemental Table 1. Conditions and Medications Used in the Exclusion Criteria**

| **Conditions** | **Medications** |
| --- | --- |
| Diabetes, myocardial infarction, stroke, symptomatic peripheral arterial disease, cancer (excluding non-melanoma skin cancer), end-stage renal disease | SGLT2:  canagliflzoin, dapagliflozin, empagliflozin, sotagliflozin, bexagliflozin, ertugliflozin  DPP4:  sitagliptin, alogliptin, linagliptin, saxagliptin  Insulin:  regular, lispro, lispro-aabc, aspart, glulisine, NPH, detemir, glargine, glargine-aglr, degludec  Sulfonylurea:  Carbutamide, chlorpropamide, glycyclamide, tolcyclamide, metahexamide, tolazamide, tolbutamide, glibenclamide, glyburide, gilbornuride, gliclazide, glipizide, gliquidone, glisoxepide, glyclopyramide, glimepiride  GLP-1 RA:  dulaglutide, tirzepatide, liraglutide, exenatide, oral semaglutide  Weight loss:  orlistat, phentermine, phentermine and topiramate, naltrexone and bupropion, setmelanotide  Combinations:  canagliflozin/metformin, dapagliflozin/metformin, empagliflozin/metformin, empagliflozin/linagliptin, ertugliflozin/sitagliptin, dapagliflozin/saxagliptin, ertugliflozin/metformin, dapagliflozin/saxagliptin/metformin, empagliflozin/linagliptin/metformin,  sitagliptin/metformin, saxagliptin/metformin, linagliptin/metformin, alogliptin/metformin, alogliptin/pioglitazone, sitagliptin/simvastatin  Insulin degludec / liraglutide, insulin glargine / lixisenatide |
| Abbreviations: DPP4, dipeptidyl peptidase-4; NPH, neutral protamine Hagedorn; GLP-1 RA, glucagon-like peptide-1 receptor agonists; SGLT2, sodium-glucose cotransporter-2. | |

**Supplemental Table 2. BMI Assignment**

| **ICD-10-CM Code** | **Classification** | **Definition** | **Assigned BMI value** |
| --- | --- | --- | --- |
| E66 | obese | Overweight or obesity | 30 |
| E66.0 | obese | Obesity due to excess calories | 35 |
| E66.01 | morbidly obese | Morbid obesity | 45 |
| E66.09 | obese | Other obesity due to excess calories | 35 |
| E66.2 | morbidly obese | Morbid obesity with alveolar hypoventilation | 45 |
| E66.3 | overweight | Overweight | 27.5 |
| E66.8 | obese | Other obesity | 35 |
| E66.9 | obese | Obesity, unspecified | 35 |
| Z68.25 | overweight | BMI 25 - 25.9 | 25.5 |
| Z68.26 | overweight | BMI 26 - 26.9 | 26.5 |
| Z68.27 | overweight | BMI 27 - 27.9 | 27.5 |
| Z68.28 | overweight | BMI 28 - 28.9 | 28.5 |
| Z68.29 | overweight | BMI 29 - 29.9 | 29.5 |
| Z68.3 | obese | BMI 30 - 39 | 35 |
| Z68.30 | obese | BMI 30 - 30.9 | 30.5 |
| Z68.31 | obese | BMI 31 - 31.9 | 31.5 |
| Z68.32 | obese | BMI 32 - 32.9 | 32.5 |
| Z68.33 | obese | BMI 33 - 33.9 | 33.5 |
| Z68.34 | obese | BMI 34 - 34.9 | 34.5 |
| Z68.35 | obese | BMI 35 - 35.9 | 35.5 |
| Z68.36 | obese | BMI 36 - 36.9 | 36.5 |
| Z68.37 | obese | BMI 37 - 37.9 | 37.5 |
| Z68.38 | obese | BMI 38 - 38.9 | 38.5 |
| Z68.39 | obese | BMI 39 - 39.9 | 39.5 |
| Z68.4 | morbidly obese | BMI 40 or greater | 45 |
| Z68.41 | morbidly obese | BMI 40 - 44.9 | 42.5 |
| Z68.42 | morbidly obese | BMI 45 - 49.9 | 47.5 |
| Z68.43 | morbidly obese | BMI 50 - 59.9 | 55 |
| Z68.44 | morbidly obese | BMI 60 - 69.9 | 65 |
| Z68.45 | morbidly obese | BMI ≥ 70 | 75 |
| Abbreviations: BMI, body mass index; ICD-10, International Classification of Diseases, 10th Revision. | | | |

**Supplemental Figure 1. Study Diagram for Semaglutide and Control Cohorts**

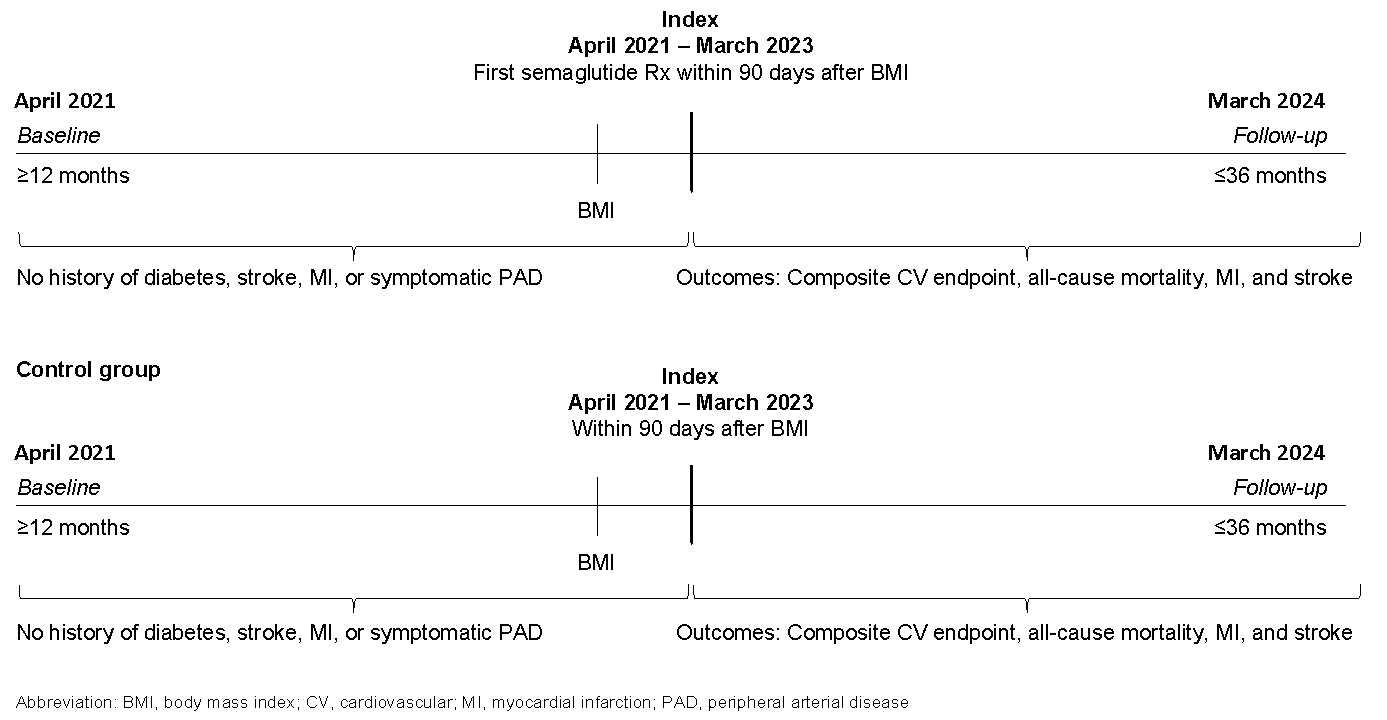

**Supplemental Figure 2. Study Flow Diagram**

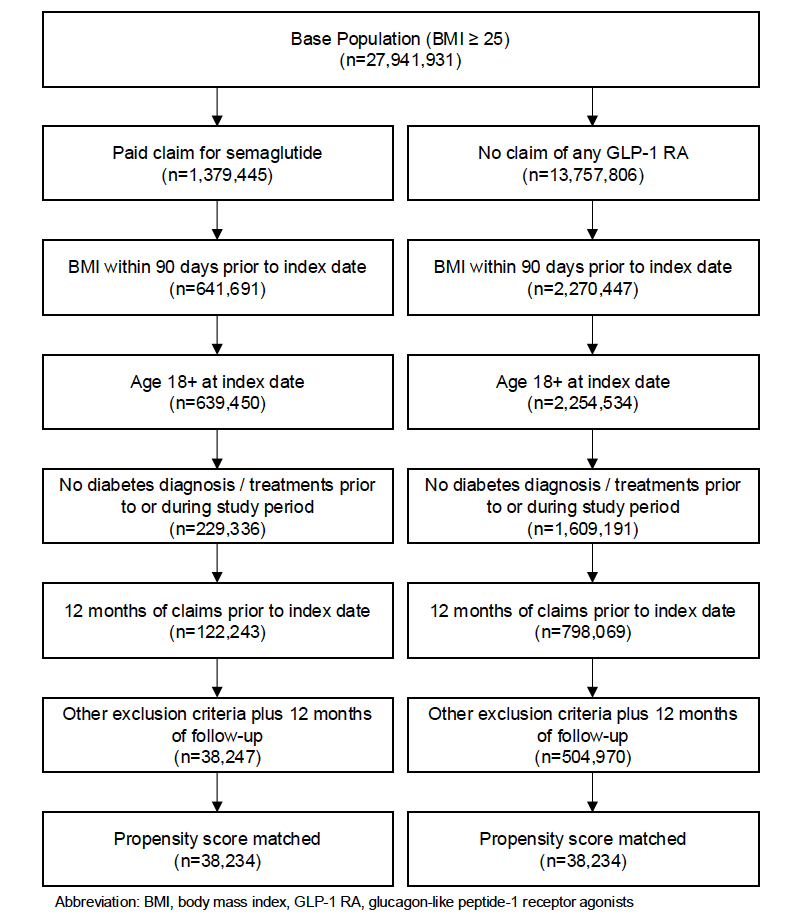
